# Unveiling the Psychological Traits of Multi-Marathoners: Insights from TIPI Personality Trait Analysis

**DOI:** 10.1101/2024.09.24.24314323

**Authors:** Leo Lundy, Richard B Reilly, Neil Fleming, Dominika Wilczyńska

## Abstract

**Objectives:** Multi-marathoners, athletes dedicated to completing 100 or more marathons, represent a unique subculture within endurance sports. This study uses the Ten Item Personality Inventory (TIPI) test to explore their psychological traits. The study aims to identify the unique personality profiles of multi-marathoners and understand their implications for participation, performance, and well-being.

**Methods:** TIPI was used to describe personality and provide valuable insights into the tendencies for certain personality traits. TIPI was conducted via an online cross-sectional survey distributed to the multi-marathon community, which received 593 responses, 56% men (n=331, average age = 53.87 years, SD = 9.91), 44% women (n=261, average age = 54.06, SD 10.56) from 22 countries. One respondent identified as another gender and was not included in the gender-based analysis. Cronbach’s Alpha and Guttman’s Lambda 6 were calculated to assess the internal consistency of the survey, and the results were statistically analysed using Mann-Whitney U tests, ANOVA Aligned Rank Transform (ART) tests, and Wilcoxon rank-sum post-hoc tests to highlight differences in emotional stability, openness, conscientiousness, extraversion, and agreeableness. Normative TIPI data from the original TIPI study served as a dataset for comparison. Additionally, Spearman’s ρ based correlation was used to explore relationships between personality traits and other related variables from the multi-marathoner community collected from a previous study.

**Results:** The findings reveal distinctive personality traits among multi-marathoners. Compared to the general population, results show multi-marathoners displayed higher levels of conscientiousness (F (1,591) = 2.42, p < 0.001 for gender), indicating strong self-discipline, organisation, and goal-oriented behaviour. They exhibited lower levels of emotional stability (F (1,591) = 5.525, p < 0.001 for age group) and openness (F (1,591) = 2.54, p < 0.001 for age group), suggesting challenges in stress management and adaptability. Following significant results from the ANOVA ART tests, the Wilcoxon rank-sum post-hoc analysis revealed a significant gender difference in agreeableness, with women exhibiting higher levels of agreeableness compared to men (W = 50809, p<0.00091). However, after applying the Bonferroni statistical correction, no significant differences were found between genders in conscientiousness or emotional stability. Additionally, there were no significant differences in personality traits across different age groups after applying the statistical correction. These findings suggest that while gender differences in agreeableness are robust, age-related differences in personality traits were not statistically significant in this study.

**Conclusion:** This study offers insights into the psychological traits of multi-marathoners, highlighting their high conscientiousness and lower emotional stability. Contrary to the initial hypotheses, no significant differences were found in openness, and age-related differences in personality traits were not statistically significant after applying the Bonferroni statistical correction. These findings suggest that while multi-marathoners possess distinct personality traits, the relationship between these traits and their engagement in endurance sports is more complex than initially anticipated. These insights can guide the development of interventions to foster resilience and sustained participation, enhancing their overall experience and success in multi-marathoning. Longitudinal studies to track changes in personality traits and explore effective psychological interventions for this unique athletic community would allow further insight.

## Introduction

Multi-marathoning, the pursuit of completing multiple marathon events, represents a unique subculture within the broader context of endurance sports. The defining achievement for many multi-marathoners is reaching the milestone of completing 100 marathons, a feat that requires extraordinary dedication, perseverance, and resilience. While a wider-ranging observational study has documented the participatory nature of multi-marathoning(1), the psychological dimensions of this endurance sport are less understood.

However, recent research on endurance athletes has increasingly revealed significant mental health concerns within this population, including heightened levels of stress, anxiety, and depression (2, 3). These findings underscore the need for a more comprehensive examination of the psychological traits that may contribute to the challenges faced by multi-marathoners, or alternatively, support their success. Therefore, this study seeks to address this gap by investigating the psychological traits of multi-marathoners, with a focus on personality characteristics. The study integrates the Big Five Personality Model (4) and Goal Setting Theory (5) to provide a theoretical framework for understanding the relationship between personality traits and participation in multi-marathon events.

Personality is often associated with the uniqueness of an individual, encompassing the characteristics that explain a person’s consistent patterns of feelings, thoughts, and behaviours (6). In a broader sense, personality can be understood as a complex integration of thoughts, emotions, and behaviours that provides direction and coherence to a person’s life. Like the body, personality consists of both structures and processes, reflecting the combined influences of genetics (nature) and environmental factors. The concept of personality also includes the temporal aspect of human functioning, as it encompasses past memories, mental representations of the present, and imaginations and expectations for the future (7). An important approach in personality psychology is trait theory, developed by, among others, Allport, Eysenck, Cattell, and Costa & McCrae. For this study, the Big Five Personality Model of Costa & McCrae were employed (4). This model categorises personality traits into five broad dimensions: openness to experience (characterised by imagination, creativity, and a preference for novelty and variety, with high scorers being more curious and open to new experiences); conscientiousness (involving traits like self-discipline, organisation, and dependability, with high conscientiousness associated with goal-oriented behaviours and careful planning); extraversion (reflecting the extent to which a person is outgoing, sociable, and energetic, with extraverts typically being more talkative and assertive, enjoying social interactions compared to introverts); agreeableness (including attributes such as trust, kindness, and empathy, with high agreeableness indicating a cooperative and compassionate nature); and neuroticism (encompassing emotional instability and the tendency to experience negative emotions such as anxiety, moodiness, and irritability, with higher levels of neuroticism associated with greater emotional reactivity, the opposite of emotional stability). This model provides a comprehensive framework for understanding these traits (6). Each of these dimensions can influence an athlete’s approach to training, goal setting, and coping with multi- marathoning. For instance, high conscientiousness might drive focused training and goal-oriented behaviour, while low emotional stability could pose challenges in managing the stress and setbacks inherent in multi-marathon participation (4).

The Goal Setting Theory (5), proposes that clear, specific, and challenging goals, combined with appropriate feedback, lead to higher performance. The theory highlights several key principles, including goal specificity, goal difficulty, and the importance of commitment, feedback, and task complexity. Specific goals direct attention and effort, challenging goals stimulate persistence and motivation, and feedback allows individuals to adjust their strategies and efforts accordingly. For multi-marathoners, these principles are particularly relevant, as the nature of their training and competition inherently involves long-term goals that require sustained commitment and adaptability. Understanding how their personality traits influence their goal-setting behaviours and resilience can offer valuable insights into the psychology factors behind sustained involvement (8).

In the context of multi-marathoning, these principles are crucial. Multi-marathoners, by the very nature of their sport, set and achieve a series of challenging goals over time, such as completing multiple marathons within a specific period. Understanding how their personality traits influence their goal-setting behaviours, resilience, and persistence can offer valuable insights into the psychological factors behind their sustained involvement. For instance, high levels of conscientiousness may drive goal planning and execution, while lower emotional stability might require more robust coping strategies to maintain commitment in the face of adversity.

Recent studies have explored the application of Goal Setting Theory in endurance sports. For example, Jackman et al. found that effective self-regulation and goal-setting were critical in achieving high performance in distance running (9). Another study by Stenling et al. reviewed the application of Goal Setting Theory across various sports and found that tailored goal-setting interventions were particularly effective in enhancing the resilience and performance of endurance athletes (10). Further studies have provided valuable insights into the relationship between personality traits and athletic performance in endurance sports. Fenta, Awoke, and Berhanu (11) examined Ethiopian long-distance runners and found that certain personality traits, such as high conscientiousness and emotional stability, were strongly associated with athletic effectiveness, underscoring the importance of these traits in sustained high performance (11). Similarly, Lopez and Sánchez (12) profiled mountain running athletes and observed that specific personality profiles, including high levels of openness and conscientiousness, contributed to success in the sport (12). Steca et al. compared the Big Five personality traits between athletes and non-athletes, finding that athletes generally scored higher in conscientiousness and extraversion, traits linked to both sports participation and success (13).

This study builds on previous observational research from a broader study that explored the participatory characteristics of the multi-marathoning community. In this broader study, through targeted survey questions, areas were explored such as motivation, diet, health, injuries, recovery and treatments (1).

By utilising TIPI, this study aims to describe specific personality traits and understand how they influence key psychological outcomes, such as goal-setting behaviours (such as achieving 100 marathon completions) and overall well-being (physical and mental health), by integrating the Big Five Personality Model with Goal Setting Theory, providing an analysis of the psychological factors that contribute to both the challenges and successes experienced by multi-marathoners. Additionally, the study explores how these personality traits may differ from those in the general population and how they might relate to sustained participation in multi-marathoning events.

Based on these theoretical frameworks and recent literature, this study hypothesises that multi- marathoners will exhibit distinct personality traits compared to the general population, notably higher conscientiousness, emotional stability and openness (4, 12). It is further hypothesised that these traits will influence outcomes such as motivation, diet, health, injuries, recovery, and treatments, which in turn may impact on their performance and well-being in the sport (1, 5).

In summary, this article aims to unveil the psychological traits of multi-marathoners, providing an understanding of how personality influences their participation and performance.

## Methods

### Study Design

A cross-sectional survey, including the Ten Item Personality Inventory (TIPI) test, was conducted online via the Qualtrics platform (14, 15), offering broad geographical reach and convenient participation. Responses were collected from multi-marathoners worldwide during the study period, from December 14, 2023, to March 31, 2024.

Inclusion criteria required respondents to be individuals who had either achieved the goal of completing 100 marathons or had set this as their running goal and were actively engaged in the sport. Exclusion criteria disqualified individuals who had minimal marathon experience and had completed fewer than two marathons.

Gatekeepers were employed to distribute the survey and were identified through an analysis of the structure of multi-marathoning at national and international levels. The study team approached all national and international multi-marathon clubs and event companies based in the UK and Ireland. All major global multi-marathon clubs and UK/Ireland-based event companies agreed to support the survey distribution through their social media channels, email groups, and newsletters, which were generally accessible to anyone interested in the sport.

### Personality Traits

Personality traits were assessed using the TIPI test (15). TIPI is a brief measure of the Big Five personality dimensions: openness, conscientiousness, extraversion, agreeableness, and emotional stability. Each dimension is measured using two items rated on a 7-point Likert scale, ranging from 1 (disagree strongly) to 7 (agree strongly). TIPI item scoring (including reverse scoring) into personality dimensions was as detailed by Gosling et al. (15). TIPI is a validated tool widely used in psychological research for reliability.

Figure 1 shows the TIPI items gathered in this study.

**Figure 1.**
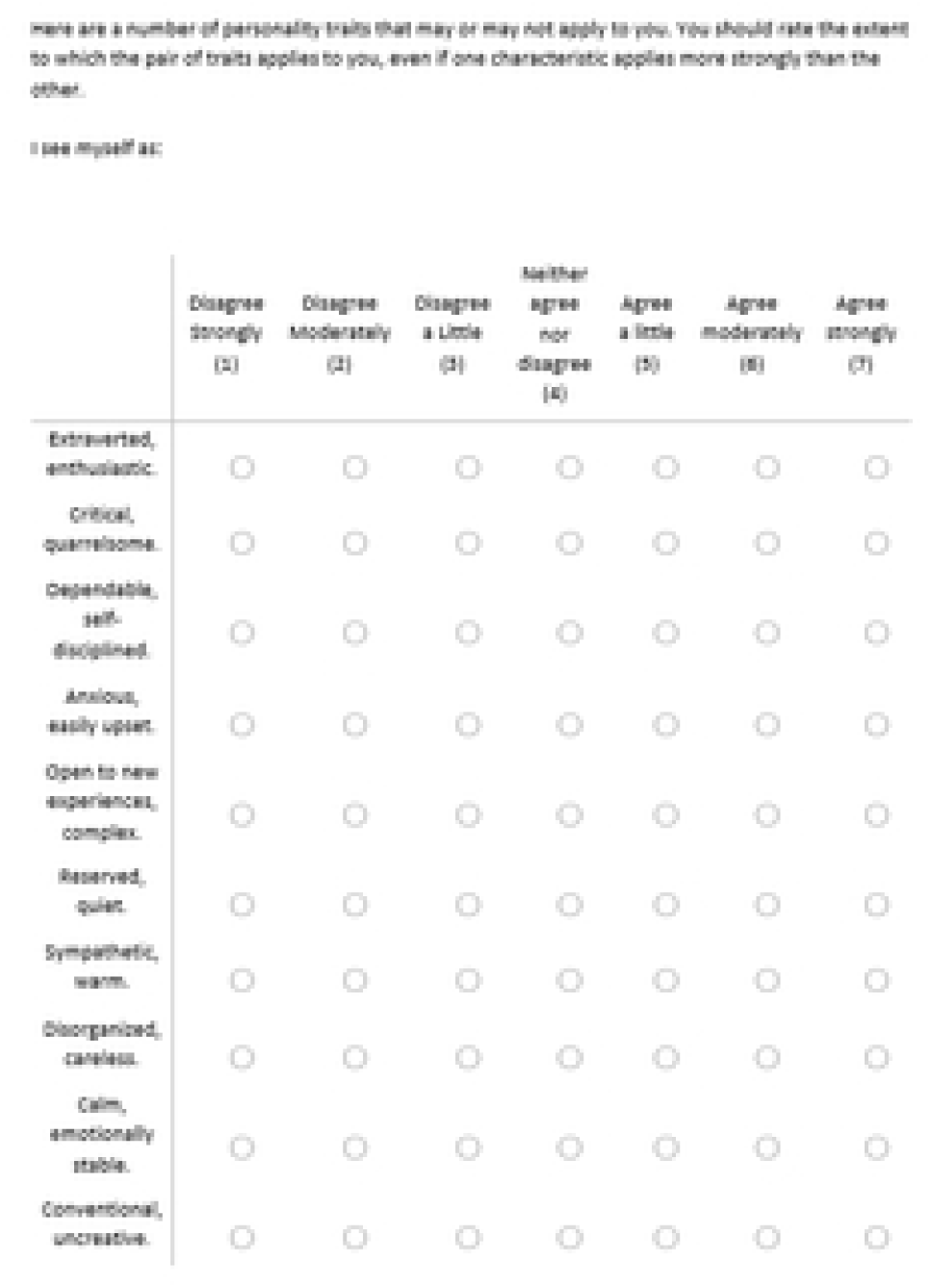
TIPI test.

### Procedure

Respondents accessed the online survey via a secure link distributed through social media channels and email lists of multi-marathon clubs and event organisations. Participation was voluntary and anonymous, with informed consent obtained from all respondents via the survey platform before their involvement, ensuring ethical standards were upheld.

### Statistical Analysis

Data were analysed using R, a statistical software package (16). Descriptive statistics were computed to summarise the demographic characteristics of the sample and the distribution of TIPI scores (15).

Normative data from the original TIPI study was used as a benchmark for comparison by age group and gender (17). This provided a large relevant normative dataset (>300,000) for the Big Five personality dimensions in the general population grouped by gender and age group.

Due to the mixed nature of the data, assumptions of normality and homoscedasticity were not applied, and statistical analysis tests were chosen that were appropriate to the mix of data under analysis.

Cronbach’s Alpha and Guttman’s Lambda 6 were calculated to assess the internal consistency and reliability of the survey (18). Mann-Whitney U tests were conducted on TIPI traits, which were ordinal in nature, to ascertain if there was statistical significance of any differences between survey and normative datasets. Mann-Whitney U tests were performed across age groups by gender (men=1, women=2) between the survey dataset and the normative dataset (13, 19). ANOVA Aligned Rank Transform (ART) Tests were used to examine the effects of gender and age group on various personality traits (20). Wilcoxon Rank-Sum Post-hoc tests identified specific pairwise comparisons with significant differences, when comparing different genders and age groups (21). Spearman’s Rank Correlation helped explore the relationships between personality traits and other various multi-marathon co-variates, where TIPI traits were correlated against relevant data from the previous observational study into multi-marathoning (1). This highlighted any significant correlations between personality traits and the various multi-marathon covariate outcomes, based on Spearman’s ρ (22).

To facilitate analysis of goal setting behaviour and correlation analysis, data from the wider-ranging observational study on multi-marathoning were analysed against TIPI results to examine goal setting behaviour and correlations across the different datasets (1). For ease of analysis, the observational study data were aggregated into correlation groups and presented in the same gender and age groups as this study data. These aggregated correlation data groups (gathered from targeted survey questions in the wider-ranging observational study) included diet, such as plant-based diets (vegan and vegetarian) and omnivorous diets (including regular, paleo, and Pesco-vegetarian); motivation, encompassing factors like achievement and recognition (e.g., competition, performance, awards, accomplishment), lifestyle and health (e.g., way of life, keeping fit), milestones and goals (e.g., running streaks, race series, race numbers, milestones), and social and enjoyment factors (e.g., travel, social life); health, covering conditions such as diagnosed heart issues, high blood pressure, balance problems, tendon and bone issues, and other chronic conditions; and injuries, which included categories such as joint and muscle injuries, common running injuries (e.g., IT band syndrome, plantar fasciitis, tendonitis, shin splints, runner’s knee), severe issues (e.g., stress fractures, frostbite, heat exhaustion), minor issues (e.g., blisters, chafing), and various treatment methods like self-care (e.g., rest, ice, compression, elevation, foam rolling, stretching, strapping), medication (e.g., pain medication), alternative therapies (e.g., acupuncture), and medical interventions (e.g., surgery). This data organisation facilitated a clearer understanding of the correlations within and across these different categories.

## Results

### Participants

A total of 593 responses were received in the cross-sectional survey of this study, 56% men (n=331, average age = 53.87 years, SD = 9.91), 44% women (n=261, average age = 54.06, SD 10.56). Only one respondent identified as another gender and was not included in the gender-based analysis. Respondents from 22 different countries across 6 continents participated, with leading countries being Ireland, UK, Germany, Finland, Italy, and the US, which is a good reflection of those countries foremost in multi-marathoning according to the multi-marathoning world rankings, thus ensuring good representation of the multi-marathon community(23). The high average marathon completion count (M:146.52 SD:196.3) among respondents further assured that the sample represented the target community. A resolution measure of ±1 year for age and ±1 marathon for the number of marathons completed was applied, acknowledging minor variances in self-reported data while maintaining the precision necessary for reliable analysis.

### Personality Traits

TIPI scores for the respondents were analysed and compared to normative data from the original TIPI study (17). Figure 2 shows the means and standard deviations (M/SD) for each TIPI survey personality trait by age group and gender and the comparisons against the TIPI normative dataset, showing differences in agreeableness, conscientiousness and emotional stability across age groups and genders. This highlights how multi-marathoners differ from the general population in terms of personality traits. It should be noted that agreeableness and emotional stability saw significant negative difference across all age groups and genders, and higher conscientiousness was only evident in women.

**Figure 2.**
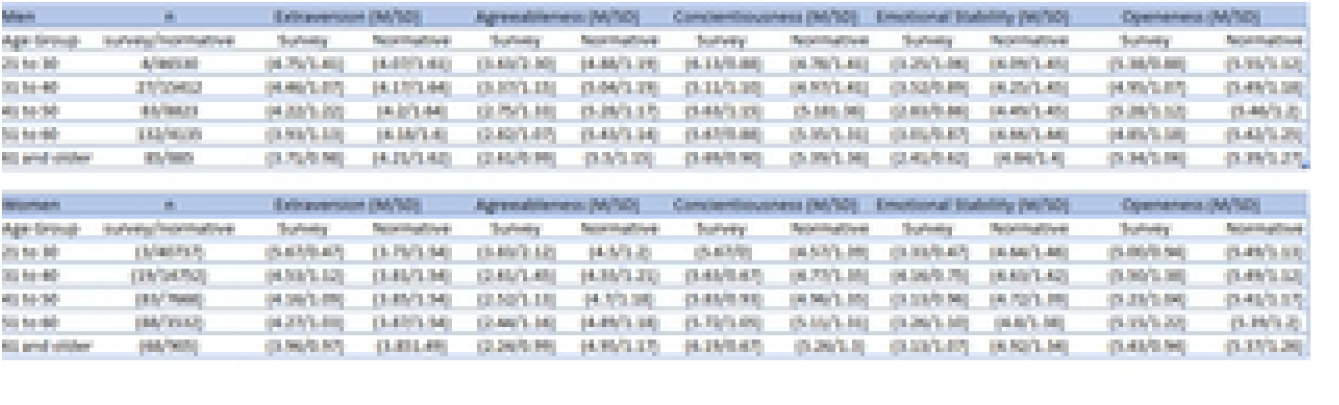
Comparison of Survey TIPI results against TIPI Normative data.

Cronbach’s alpha (0.42) and Guttman’s Lambda 6 (0.40) revealed lower reliability compared to longer personality assessments (15, 18, 24, 25). However, this level of reliability is considered moderate within the context of ultra-brief scales like the TIPI, which assesses five traits using only 10 items. The trade-off between brevity and internal consistency is well-documented for such scales, where the focus is on efficiency rather than high internal consistency (15, 24, 25). This trade-off was accepted by the research team for the purposes of this study.

Mann-Whitney U tests showed agreeableness, conscientiousness, and emotional stability had statistically significant differences with alpha = 0.05 and a Bonferroni statistical correction of 50 (5 traits by 10 age groups), with p <0.001 (significant), P< 0.0002 (very significant), p <0.00002 (extremely significant).

Table 1 shows the p-values of Mann Whitney U tests for all traits of TIPI when compared against the normative dataset. In all the older age groups agreeableness, conscientiousness (women only), and emotional stability saw significant differences to the normative dataset.

**Table 1.** P-value results for Mann Whitney U tests by age group and gender (men =1, women =2).

| Age Category | Gender | Extraversion | Agreeableness | Conscientiousness | Emotional Stability | Openness |
| --- | --- | --- | --- | --- | --- | --- |
| 21-30 | 1 | 0.118 | 0.109 | 0.343 | 0.886 | 0.3223 |
| 31-40 | 1 | 0.471 | <0.00002 | 0.61 | 0.069 | 0.0051 |
| 41-50 | 1 | 1 | <0.00002 | 0.722 | <0.00002 | 0.4515 |
| 51-60 | 1 | 0.003 | <0.00002 | 0.001 | <0.00002 | 0.0001 |
| 61 and older | 1 | 0.17 | <0.00002 | 0.001 | <0.00002 | 0.9231 |
| 21-30 | 2 | 0.2 | 0.396 | 0.398 | 0.202 | 1 |
| 31-40 | 2 | 0.204 | 0.002 | 0.031 | 0.039 | 0.314 |
| 41-50 | 2 | 0.064 | <0.00002 | <0.00002 | <0.00002 | 0.194 |
| 51-60 | 2 | 0.148 | <0.00002 | <0.00002 | <0.00002 | 0.085 |
| 61 and older | 2 | 0.401 | <0.00002 | <0.00002 | <0.00002 | 0.685 |

### ANOVA Aligned Rank Transform Results

**ANOVA Aligned Rank Transform (ART)** tests were conducted on TIPI traits to examine the effects of Gender and Age groups and their interaction. Alpha = 0.05 with p <0.05 (significant), P< 0.001 (very significant), p <0.0001 (extremely significant).

Table 2. shows the results of these ANOVA ART tests. Extraversion, agreeableness, emotional stability, and openness showed significant differences primarily due to age group. In the case of conscientiousness, there is a significant effect of both gender and age group. No significant interaction effects between gender and age group were found for any of the traits.

**Table 2.**
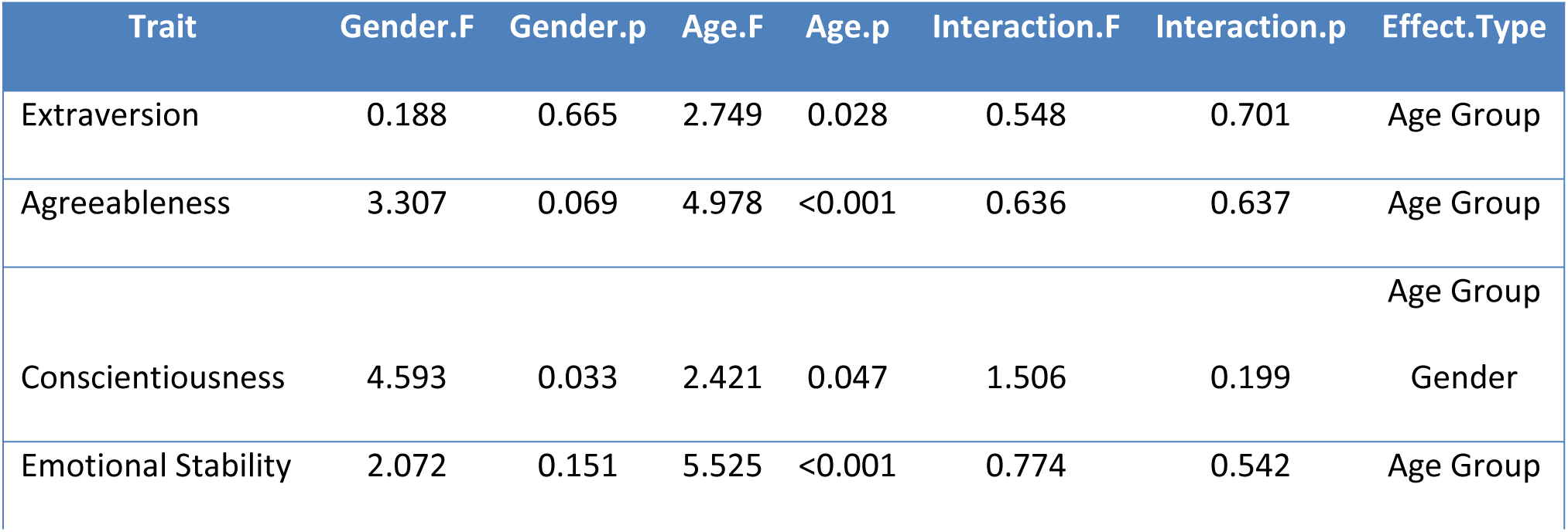
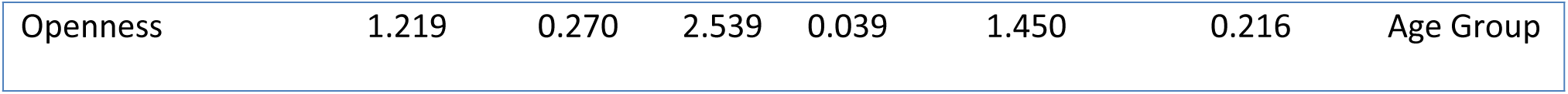
ANOVA ART results examining the effects of Gender and Age Group on Traits.

As significant results were observed from the ANOVA ART tests, the Wilcoxon rank-sum post-hoc analysis was applied to identify specific pairwise comparisons with significant differences in personality traits. This allowed the differences between gender, age groups, W statistic and p-value for each significant comparison to be detailed at an alpha level of 0.05. As 55 pairwise post hoc tests were performed (5 traits by (10 age groups plus gender)) a Bonferroni correction of 55 was applied to alpha with the resultant p<0.00091 (significant), p<0.000018 (very significant), and p<0.0000018 (extremely significant).

The results of the Wilcoxon rank-sum post-hoc tests are shown in Table 3 and once the Bonferroni statistical correction is considered there is only a significant difference found in agreeableness where women demonstrate higher levels of agreeableness to men.

**Table 3.** Wilcoxon rank-sum post-hoc tests by Gender (men =1, women = 2) and Age-Group.

| Trait | Effect Type | W statistic | p-value |
| --- | --- | --- | --- |
| Agreeableness | Gender: 1 vs 2 | 50809 | <0.0009 |
| Agreeableness | Age Group: 61 and older vs 21-30 | 191 | 0.004 |
| Conscientiousness | Gender: 1 vs 2 | 36638.5 | 0.001 |
| Emotional Stability | Gender: 1 vs 2 | 36956.5 | 0.002 |

These findings highlight the complex impact of demographic factors on personality dimensions and underscore the importance of considering personality traits when developing support mechanisms and interventions aimed at enhancing the well-being and performance of multi- marathoners.

### Goal Setting Behaviour and Correlations compared with data from previous broader observational study

As part of the previous broader observational study into participation characteristics of multi- marathoners, its cross-sectional survey had a total of 830 responses: 60.69% men (n=502, average age = 51.6 years, SD=9.96) and 39.3% women (n=326, average age = 48.83 years, SD=9.15). A small proportion (0.1%) identified as gender variant/non-conforming and were excluded from gender-based analysis. Data including diet, motivation, injury, recovery, treatment, and health were gathered using targeted survey questions. These questions are detailed in the reference for further consultation (1).

While goal setting is inherent in multi-marathoners (achieving 100 marathon completions) goal setting behaviour was further analysed in the motivation related survey questions of the broader observational study, which including questions covering achievement, recognition, milestones, and goals (1).

Results from this broader observational study showed most motivations identified were intrinsic, with maintaining fitness and health being the most common (18.32%). This was followed closely by a sense of accomplishment (16.26%) and multi-marathoning as a way of life (13.62%). Social engagement also played a significant role (11.53%), alongside motivations such as travel (8.82%) and reaching specific milestones (8.3%).

For men with fewer than 100 marathon completions, motivations were largely centred on continuous improvement and competition. However, among men with over 100 completions, these motivations shifted, with travel and social engagement becoming the primary driving factors. In contrast, women prioritised social life across both groups, but for those with fewer than 100 marathon completions, reaching milestones was particularly important as was travel and social engagement.

In terms of extrinsic motivations, many multi-marathoners were members of multi-marathon awards clubs. Notably, 45% of respondents identified achieving 100 marathon completions and earning the associated 100-marathon award from their national club as their most significant goal.

To extend the analysis with this previous multi-marathon wider-ranging study, correlations using Spearman’s ρ were applied between TIPI traits from this study and all data from the previous multi-marathon observational study into participation in multi-marathoning including diet, motivation, injury, recovery, treatment, and health. (1, 22). Spearman’s ρ is categorised into significance levels 0-0.19: Very Weak, 0.2-0.39: Weak, 0.4-0.59: Moderate, 0.6-0.79: Strong, 0.8-1: Very Strong. The corelation analysis found 9 occurrences of Strong or Very Strong significance levels across correlations.

Table 4. details the results from this correlation analysis where the correlation had a strong or very strong significance between the TIPI traits from this study and all data from the previous broader study.

**Table 4.** Strong or Very Strong significance levels (Spearman’s ρ) of correlations of TIPI TRAITS.

| TIPI Trait | Correlated variables | Spearman's $\rho$ |
| --- | --- | --- |
| Extraversion | Health: High Blood Pressure | -0.62 |
| Agreeableness | Health: Chronic illness | -0.67 |
| Emotional Stability | Health: High Blood Pressure | -0.82 |
| Emotional Stability | Health: Balance | 0.7 |
| Agreeableness | Motivation: Social and Enjoyment | -0.71 |
| Extraversion | Injury: Minor Issue | 0.64 |
| Extraversion | Injury: Running Injury | -0.69 |
| Emotional Stability | Injury: Minor Issue | 0.66 |
| Emotional Stability | Injury: Running Injury | -0.78 |

Extraversion is negatively correlated with blood pressure (ρ = -0.62) and running injuries (ρ = -0.69), indicating that more extraverted individuals tend to have lower blood pressure and are prone to fewer running-related injuries. Additionally, higher extraversion is positively correlated with a greater incidence of minor skin issues (ρ = 0.64), suggesting that more extraverted individuals may be more prone to minor skin problems.

Agreeableness **i**s negatively correlated with chronic illness (ρ = -0.67), meaning that more cooperative and empathetic individuals tend to report fewer chronic health conditions. Interestingly, agreeableness is also negatively correlated with motivation related to social and enjoyment factors (ρ = -0.71), possibly indicating that individuals who score high in agreeableness may be less driven by social motivations in multi-marathoning, which could seem counterintuitive but warrants further exploration.

Emotional stability is strongly negatively correlated with high blood pressure (ρ = -0.82) and a lower incidence of running injuries (ρ = -0.78), suggesting that more emotionally stable individuals tend to have better physical health outcomes in these areas. Additionally, emotional stability is positively correlated with better physical balance (ρ = 0.70) and fewer minor skin issues (ρ = 0.66), indicating that emotionally stable individuals may also have better physical coordination and fewer minor injuries.

These results show that while a strong or very strong correlation can be made between personality traits and some hypothesised outcomes (such as motivation, injury, and health), other variables such as diet, recovery, and treatment showed moderate or weak correlations to personality traits contrary to initial expectations.

## Discussion

### Principal Findings

This study aimed to investigate the psychological traits of multi-marathoners, focusing on their personality profiles as assessed by the Ten Item Personality Inventory (TIPI) (15, 17). The findings reveal significant differences in specific personality dimensions between multi- marathoners and the general population. Specifically, multi-marathoners exhibited higher levels of conscientiousness and lower levels of emotional stability. However, contrary to expectations, no significant differences were observed in openness, and age-related differences in personality traits were not statistically significant after applying the Bonferroni correction. Additionally, the study found a significant gender difference in agreeableness, with women displaying higher levels compared to men. However, no significant gender differences were observed in conscientiousness or emotional stability after applying the Bonferroni correction.

### Interpretation of Results

The higher levels of conscientiousness observed among women suggest that they possess strong self-discipline, organisation, and goal-oriented behaviours (6). These traits are essential for the physical demands and persistence required to complete multiple marathons. Conscientious individuals are likely to be methodical in their preparation, setting and adhering to long-term goals, which aligns well with the demands of multi-marathoning.

Conversely, the lower levels of emotional stability indicate that multi-marathoners may face challenges in managing stress and regulating emotions. Emotional stability, often referred to as neuroticism in the Big Five Personality Theory, relates to an individual’s tendency to experience negative emotions such as anxiety, moodiness, and irritability. The demands of continuous multi-marathoning can be significant stressors, and lower emotional stability may exacerbate these challenges, potentially leading to higher levels of psychological distress (4).

The absence of significant differences in openness does not support the initial hypothesis that multi-marathoners would exhibit higher levels of this trait due to the adventurous nature of the sport. This might suggest that while multi-marathoners engage in a physically demanding and challenging activity, they may prefer the structured and predictable aspects of their event routines.

### Gender and Age Effects

The analysis indicated a significant gender difference in agreeableness, with women demonstrating higher levels than men. This suggests that while women may excel in cooperative and compassionate behaviours, the anticipated gender differences in conscientiousness and emotional stability did not hold after statistical correction. Additionally, the expected age-related trends were not statistically significant after applying the Bonferroni correction, indicating that older age groups did not exhibit the anticipated lower levels of agreeableness and emotional stability or higher levels of openness. This suggests that the interaction between age and personality traits might be more complex than initially hypothesised.

### Theoretical Implications

The findings of this study can be interpreted through the frameworks of the Big Five Personality Theory and Goal Setting Theory. The Big Five Personality Theory provides a robust framework for understanding how individual differences in personality traits influence behaviour (6). In this context, high conscientiousness and low emotional stability among multi- marathoners highlight the dual aspects of persistence and vulnerability within this group.

Goal Setting Theory complements these insights by emphasising the importance of clear, challenging goals and sustained commitment to achieving multi-marathon goals. The strong goal-oriented behaviour associated with high conscientiousness supports the notion that multi- marathoners are particularly adept at setting and pursuing long-term objectives, which is crucial for their sport (5).

### Practical Implications

Understanding the personality traits of multi-marathoners has practical implications for designing interventions aimed at optimising their well-being and performance. Given the high levels of conscientiousness among women, support programmes should leverage this trait by providing structured gender-specific training plans and goal-setting workshops that align with their methodical approach to marathon preparation.

For those with lower emotional stability, psychological support mechanisms such as stress management training, resilience building, and access to mental health resources are essential. These interventions can help multi-marathoners better manage the psychological demands of their sport and mitigate the risks associated with lower emotional stability (26).

Since the findings didn’t show a strong link between openness (a personality trait likely involving adaptability, creativity, or willingness to experience new things) and performance, support programs may not need to focus heavily on fostering these qualities. However, it is still important to keep athletes motivated and engaged using diverse training methods.

### Intervention

The insights gained from this research can inform interventions designed to address and enhance the psychological resilience and overall well-being of multi-marathoners, ensuring they achieve their race goals while also maintaining a balanced and healthy psychological state.

### Future Research Directions

Future research should explore longitudinal changes in personality traits among multi- marathoners to understand how their personality profiles evolve with continued participation in the sport. Investigating the impact of specific interventions designed to enhance psychological resilience and well-being can provide evidence-based strategies for supporting multi-marathoners (27).

Moreover, expanding the diversity of study samples to include a broader range of cultural, socioeconomic, and geographic backgrounds can enhance the generalisability of the findings and provide a more nuanced understanding of the psychological challenges faced by multi- marathoners globally.

### Ethical Considerations

The study was conducted in accordance with the Declaration of Helsinki and received ethical approval from Trinity College Dublin’s Faculty of Health Sciences Research Ethics Committee (FREC) Number 231005.

### Limitations

While the study aimed to maintain respondent anonymity and reduce potential biases, several limitations should be noted:

#### Selection Bias

The survey distribution through gatekeepers in closed social media groups might introduce selection bias, affecting the generalisability of the results.

#### Response Bias

Using anonymous self-reported data may lead to response bias, where participants provide socially acceptable or inaccurate answers.

#### Unpublished data

Use of unpublished TIPI norms provided by Gosling et al. for comparison purposes (17). While these norms are widely referenced in personality research, their unpublished status means that they may not have been subjected to the same rigorous peer review as published data. This could introduce some uncertainty regarding the accuracy or representativeness of the normative data used as a benchmark in this study.

#### Lack of Insights from Dropouts

The survey did not include insights from individuals who stopped pursuing multi-marathoning, limiting understanding of factors influencing participation and retention.

Despite these limitations, the study offers important insights into different aspects of multi- marathoning, setting the stage for future research, which should consider longitudinal designs and objective measures of psychological health to build on the findings of this study.

### Research/Policy implications

Until now, the absence of peer-reviewed research has resulted in a lack of understanding of nature of the sport.

Multi-marathon participants and those involved in multi-marathon governance, in the provision of multi-marathon events, or health professionals, may utilise the recommendations given in this study to better plan their contribution to the sport and its overall safety, policies and organisation.

## Conclusion

This study highlights distinctive personality profiles among multi-marathoners, characterised by high conscientiousness and lower emotional stability. These traits underline the importance of self-discipline in meeting the demands of multi-marathoning while also pointing to potential psychological challenges in stress management. Contrary to initial expectations, no significant differences were found in openness or in the interaction between age and personality traits, suggesting that the relationship between personality and multi-marathoning may be more complex than previously thought.

By applying the Big Five Personality Theory and Goal Setting Theory, the research provides insights that can guide the development of targeted interventions to support the well-being and performance of multi-marathoners. Practical implications include the need for structured training programs and stress management resources to help athletes maintain both their physical and mental health.

Future research should explore longitudinal changes in personality traits and test the effectiveness of psychological interventions. Additionally, broadening the diversity of study samples will enhance the generalisability of these findings across the global multi-marathon community.

In conclusion, while personality traits such as conscientiousness and emotional stability play a significant role in shaping the experiences and outcomes of multi-marathoners, the anticipated effects of openness and age on personality were not confirmed, offering a refined perspective for sports psychology and endurance sports research.

## Abbreviation

FREC: Faculty of Health Sciences Research Ethics Committee
TIPI: Ten Item Personality Inventory

## Acknowledgments

The authors of this study would like to offer thanks to all study participants for their contribution to this research.

## Patient consent for publication

Not applicable

## Provenance and peer review

Not commissioned, peer-reviewed

## Data availability statement

Data are available in line with PLoS ONE policy

## Notes on Authors

**Leo Lundy,** BSc, is a PhD candidate at the School of Engineering at Trinity College, The University of Dublin.

**Richard Reilly,** PhD is Professor of Neural Engineering at Trinity College, The University of Dublin, a joint position between the School of Medicine and School of Engineering. He is Director and a principal investigator of the Trinity Centre for Bioengineering (TCBE) and a principal investigator at the Trinity College Institute of Neuroscience (TCIN). His research focuses on high-density electrophysiological and neuroimaging-based analysis of sensory and cognitive processing for clinical applications.

**Neil Fleming.** PhD is an assistant professor and director of the Human Performance Laboratory, Trinity College Dublin. Involved in neuromuscular physiology research within the School of Medicine.

**Dominika Wilczyńska**, PhD, Associate Professor, Faculty of Social and Humanities, University WSB Merito, 80-226 Gdańsk, Poland. Primary research focus is on positive sports psychology, to enhance well-being and reduce burnout in both young and adult athletes.

